# Microvascular Responses to Common Endothelial Stimuli Are Not Related in Humans

**DOI:** 10.64898/2026.07.02.26357129

**Authors:** Kelsey S. Schwartz, Madison G. Evering, Claire E. Goebel, Jody Greaney, Anna E. Stanhewicz

## Abstract

**Background:** Cutaneous microvascular responses to local heating and acetylcholine perfusion are widely used to assess nitric oxide (NO)-mediated endothelium-dependent dilation in human health and disease. Despite the increasingly common usage of these approaches, no studies have directly compared responses to these stimuli within individuals. Therefore, we assessed endothelium- and NO-dependent dilation in 80 young adults (40 males/40 females; 22±3 years) to determine the extent to which microvascular responses to these endothelium-dependent stimuli are comparable within an individual.

**Methods:** We examined cutaneous vascular conductance responses to (1) standardized local heating protocols to 39°C and 42°C, and (2) graded infusions of acetylcholine (10^-10^-10^-1^ M) alone or with 15 mM N^G^-nitro-l-arginine methyl ester (L-NAME; NO synthase inhibitor). Endothelium- and NO-dependent dilation were calculated and expressed in multiple ways based on commonly published analyses to allow for a thorough comparison within and between each stimulus.

**Results:** Local heating-induced endothelium- and NO-dependent dilation were lower at 39°C compared with 42°C (P<0.001). The magnitude of local heating-induced endothelium-dependent dilation was significantly related to the NO-dependent contribution of that response at 39°C (R^2^= 0.79) and 42°C (R^2^= 0.56). Local heating-induced NO-dependent dilation at 39°C was not related to that at 42°C (P>0.05). Acetylcholine-induced endothelium- and NO-dependent dilation were not related to local heating-induced responses (all P>0.05).

**Conclusions:** These data demonstrate that while local heating and acetylcholine perfusion produce robust endothelium- and NO-dependent cutaneous vasodilation, these responses are not comparable within an individual.

**Clinical Trial Registration:** URL: https://www.clinicaltrials.gov/ Unique identifier: NCT06499844

**Novelty and Relevance:** *What Is New?:*

- Local thermal heating and acetylcholine perfusion each produce robust endothelium- and nitric oxide-dependent cutaneous vasodilation. However, these responses are not directly comparable within an individual.

*What Is Relevant?:*

- Changes in microvascular function precede and predict adverse changes in conduit vessels and are reversible before the onset of overt vascular disease and target organ damage.

*Clinical/Pathophysiological Implications?:*

- These approaches are widely used by investigators to interrogate mechanisms of microvascular function and dysfunction in humans. Selection of the appropriate approach should be based on the specific research question and pathophysiological mechanism under investigation.

## INTRODUCTION

The human cutaneous circulation is an easily accessible, representative vascular bed for examining mechanisms of microvascular function and dysfunction in vivo^1,2^. Coupled with intradermal microdialysis for the acute, local delivery of pharmacological agents directly to the dermis, cutaneous vascular responses to physiological and pharmacological stimuli allow for the mechanistic examination of microvascular signaling pathways in humans. We and others have shown that cutaneous vascular endothelial responses are reduced in populations at higher risk for future cardiovascular disease (CVD)^3–5^ or with overt clinical disease^6–9^. Further, changes in the microvasculature precede and predict adverse changes in conduit vessels^10,11^ and vascular endothelial dysfunction is reversible with lifestyle or pharmacological interventions prior to the progression of overt disease^12,13^. Thus, the cutaneous circulation is a valid and sensitive bioassay for interrogating mechanisms of microvascular endothelial function and evaluating interventional approaches in humans.

A variety of physiological and pharmacological perturbations have been used to assess cutaneous microvascular endothelium- and nitric oxide (NO)-dependent dilation, including local heating of the skin and intradermal delivery of exogenous acetylcholine. Local heating-induced vasodilation, often to 39°C or 42°C, evokes a physiological endothelium-dependent plateau in skin blood flow that is primarily NO-mediated^14,15^. Although local heating to 42°C has historically been the more standard approach, local heating to 39°C has become increasingly common following a report suggesting a greater contribution of NO to the plateau response at 39°C (∼90%)^14^ compared to that at 42°C (∼70%)^15,16^. More recently, perfusion of exogenous acetylcholine in a dose-dependent manner has been used to examine endothelium-dependent dilation, although it induces a response that is less dependent on NO (∼30-40%)^17,18^. Several studies have utilized local heating and/or acetylcholine perfusion to quantify and compare endothelium- and NO-dependent dilation in healthy and clinical populations. These studies consistently report reductions in microvascular endothelial function in high risk and clinical CVD populations across techniques^3,9,17,19–25^. However, no studies have examined if endothelium- and NO-dependent dilation responses to these three commonly used stimuli are related and/or directly comparable when tested within an individual, limiting comparisons across studies that utilize these conceptually similar, yet technically distinct, approaches.

Therefore, the purpose of this study was to investigate whether the relative magnitude of cutaneous microvascular endothelium- and NO-dependent dilation in response to local heating and acetylcholine perfusion are comparable when assessed simultaneously within an individual (i.e., an individual with blunted responses to 39°C would also exhibit blunted responses to both 42°C and acetylcholine). Within local heating stimuli, we hypothesized that the magnitude of endothelium- and NO-dependent dilation would be greater during 42°C compared with 39°C, but these responses would be correlated within an individual. Between physiological and pharmacological stimuli, we hypothesized that the NO-dependent contribution to endothelium-dependent dilation would be greater in response to local heating (39°C and 42°C) than to acetylcholine, and that local heating- and acetylcholine-induced endothelial responses would not be correlated within an individual.

## METHODS

### Ethical Approval

The data that support the findings of this study are available from the corresponding author upon reasonable request. All protocols were approved by the University of Iowa Institutional Review Board (IRB; no. 202309383) and the University of Delaware IRB (no. 2190901). Written and verbal informed consent was obtained before study enrollment in accordance with the Declaration of Helsinki. Eighty adults aged 18-30 participated in the study. Forty participants (20 males/20 females) were enrolled at each site. We refer the reader to our companion paper (Evering et al., 2026) for a thorough analysis of sex differences in this cohort.

See Supplemental Material for detailed methods.

### Microvascular Reactivity Measures

Four intradermal microdialysis fibers (CMA 31 Linear Microdialysis Probe, CMA Microdialysis, Holliston, MA) were placed in the left ventral forearm for the local delivery of pharmacological agents. Three protocols to assess endothelium- and NO-dependent dilation were simultaneously performed as described below.

### Cutaneous Local Heating Protocol

Two microdialysis fibers were randomly selected to receive lactated Ringer’s (control) to undergo a local heating protocol to either 39°C or 42°C. Following baseline measurements, endothelium-dependent dilation was assessed using a standardized local heating protocol to 39°C or 42°C (0.1°C·s^-1^)^3,16,26,27^. Once plateaued, both sites were perfused with 15 mM L-NAME (NO synthase inhibitor) to determine site-specific NO-dependent dilation.

### Cutaneous Vasodilator Responsiveness to Acetylcholine

The 2 remaining microdialysis fibers were randomly selected to receive either lactated Ringer’s (control) or 15 mM L-NAME. After baseline, ascending concentrations of acetylcholine (10^-10^-10^-1^ M; United States Pharmacopeia, Rockville, MD) alone (control) or with L-NAME were perfused sequentially for 5 minutes each^5,21,28,29^.

### Maximal Skin Blood Flow

Following completion of the above protocols, all microdialysis fibers were perfused with 28 mM sodium nitroprusside (United States Pharmacopeia) and local heaters were increased to 43°C to elicit site-specific maximal dilation^3,14,16,19,20,26,27^.

### Analytical Procedures

Absolute cutaneous vascular conductance was calculated (CVC=laser-Doppler flux/mean arterial pressure) and normalized to a percentage of site-specific maximum (relative CVC, %max) for analysis^3,5,9,14–21,26–30^.

### Quantification of local heating-induced responses

Local heating (LH)-induced endothelium-dependent dilation was quantified during a stable 3-5 min plateau in skin blood flow^3,26,27^. The primary quantification of NO-dependent dilation was calculated as the difference between the LH- and L-NAME-induced plateaus [NO {Δ%max} = LH plateau–L-NAME plateau]. Secondary analysis of NO-dependent dilation were calculated as (1) a percentage of the LH plateau [NO {%plateau} = (LH plateau–L-NAME plateau)/(LH plateau)×100] and (2) a percentage of the LH plateau above baseline [NO {%plateau_BL_} = [(LH plateau−baseline)−(L-NAME plateau−baseline)]/(LH plateau−baseline)×100], according to recent recommendations^16,31^.

### Quantification of acetylcholine-induced responses

Individual acetylcholine-induced endothelium-dependent dilation was quantified from the control site as: (1) area under the curve (AUC; arbitrary units, a.u.), (2) predicted CVC (%max) at individual logEC_50_ and logEC_90_ (see *Pharmacological Curve Modeling* in Supplemental Materials); and (3) peak CVC (%max), defined as the maximum vasodilatory response during acetylcholine perfusion. For each approach, NO-dependent dilation was quantified as: (1) the difference in AUC between sites [NO {Δa.u.} = control AUC–L-NAME AUC]; (2) the difference between the predicted CVC (%max) at the logEC_50_ or logEC_90_ between sites [NO {Δ%NOpredicted} = predicted control CVC–predicted L-NAME CVC at the same dose]; (3) the difference in AUC between sites as a percentage of the control site AUC [NO {%AUC} = (control AUC–L-NAME AUC)/(control AUC)×100]; and (4) the difference between sites at the acetylcholine dose that elicited the peak CVC response at the control site [NO {Δ%max} = peak control CVC–L-NAME CVC at the same dose]. NO-dependent dilation quantified as the difference in AUC (Δa.u) and the difference in predicted CVC (%NOpredicted) were employed as primary analyses.

One-way repeated measures ANOVAs were used to assess differences between local heating protocols (39°C vs. 42°C) for (1) local heating phases and (2) NO-dependent dilation calculated using each mathematical approach. A two-way repeated measures ANOVA was used to detect differences between acetylcholine sites. A paired t-test was used to compare NO-dependent dilation between logEC_50_ and logEC_90_. Simple linear regression analyses were used to evaluate associations between endothelium- and NO-dependent dilation between 39°C and 42°C and within local heating and acetylcholine protocols. Pearson’s correlation analyses were used to assess relations between vascular responses to local heating and acetylcholine.

## RESULTS

Participant characteristics are presented in Supplemental Table 1. Twenty-one females reported hormonal contraception use (12 oral pill, 5 intrauterine device, 3 implant, 1 patch). The average self-reported day of menstruation during the experimental visit was day 14 (range 2-33).

### Microvascular endothelium- and NO-dependent dilation responses are greater during local heating to 42°C compared with 39°C

Relative CVC (%max) for each phase of both local heating protocols are presented in Figure 1. Baseline and L-NAME plateau responses were not significantly different between local heating to 39°C and 42°C. However, the magnitude of the axon-mediated peak and the local heating plateau responses were reduced at 39°C compared with 42°C (both P<0.001). Figure 2 presents local heating-induced NO-dependent dilation. NO-dependent dilation expressed as the difference between the local heating and L-NAME plateaus (Δ%max) and as a percentage of the local heating plateau attributable to NO (%plateau) were both lower at 39°C compared with that at 42°C (both P<0.001; Figure 2A-B). There was no difference in NO-dependent dilation between protocols when expressed as a percentage of the local heating plateau above baseline (%plateau_BL_) (P=0.94; Figure 2C). There were no significant differences in absolute CVC between sites (Supplemental Table 2).

**Figure 1.**
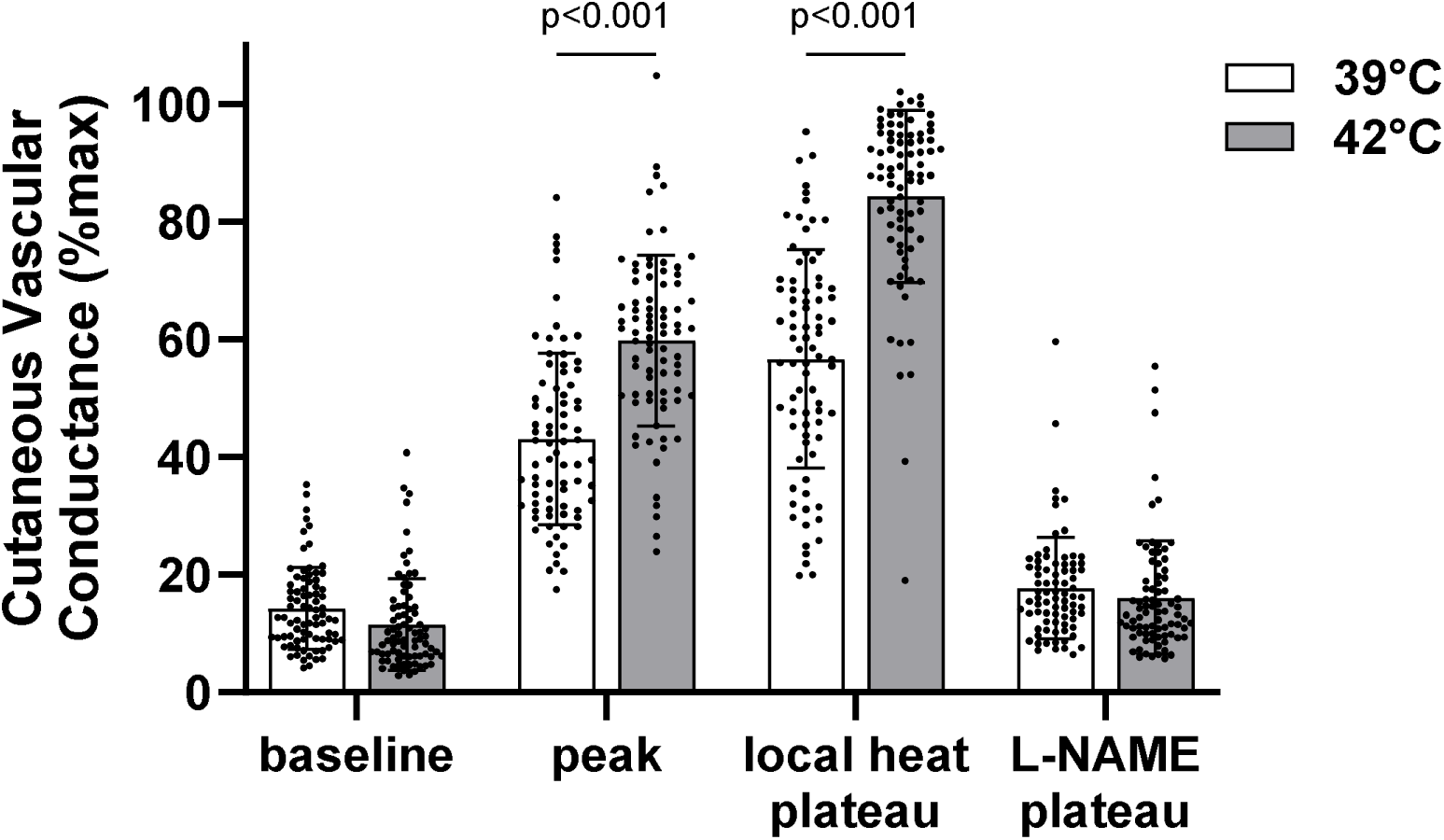
Microvascular responses (cutaneous vascular conductance, %max) during local heating of the skin to 39°C and 42°C. Data are shown for each protocol at baseline, the axon-mediated peak (peak), the endothelium-dependent plateau during local heating (local heat plateau), and the nitric oxide synthase inhibition plateau [L-NAME (N^G^-nitro-l-arginine methyl ester) plateau]. P-values determined by one-way repeated measures ANOVA with Tukey post hoc.

**Figure 2.**
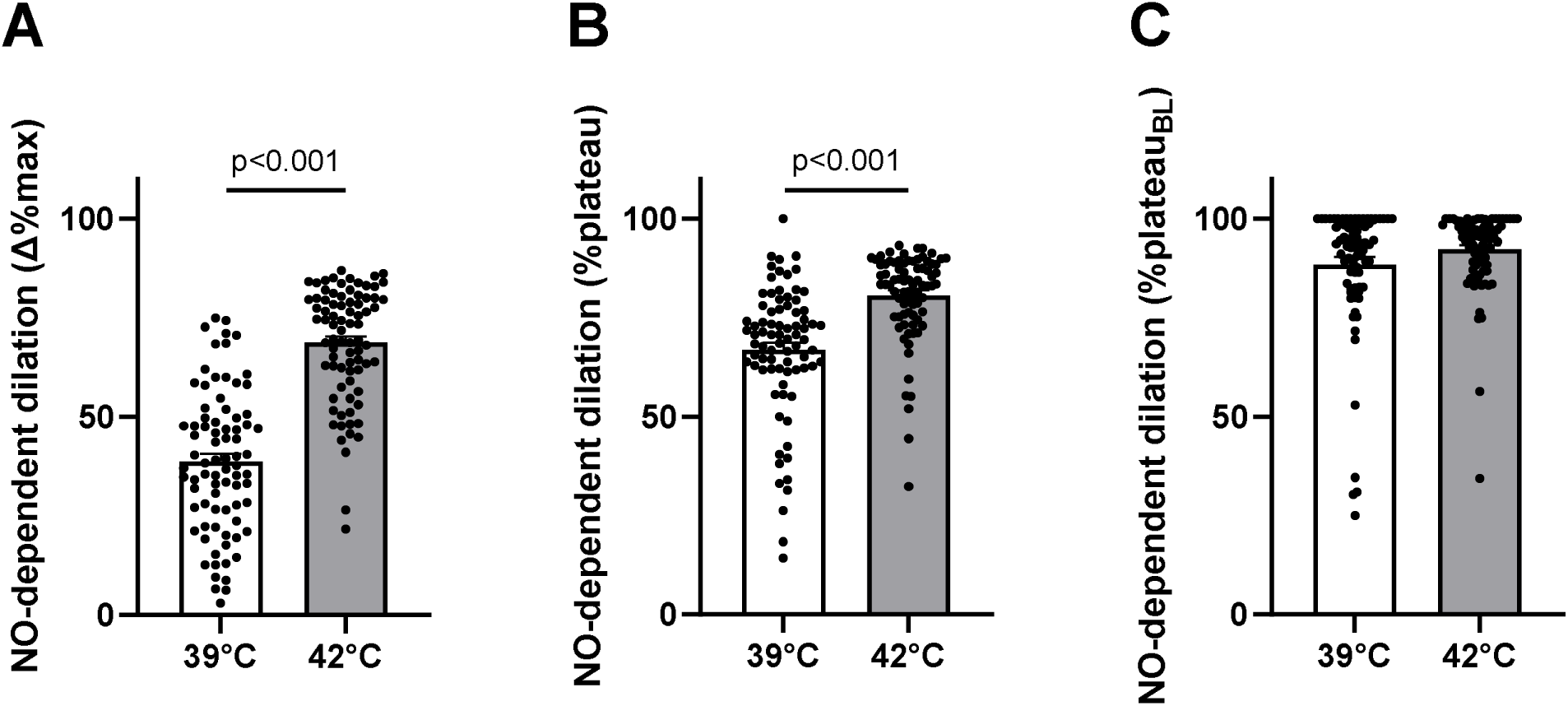
Nitric oxide (NO)-dependent vasodilation responses during local heating to 39°C and 42°C. NO-dependent dilation expressed as the difference between the local heat and L-NAME (N^G^-nitro-l-arginine methyl ester) plateau (A; Δ%max), a percentage of the local heating plateau (B; %plateau), and a percentage of the local heating plateau above baseline (%plateau_BL_). See *Analytical Procedures* for calculations. P-values determined by one-way repeated measures ANOVA with Tukey post hoc.

### Associations between endothelium- and NO-dependent dilation responses to local heating are dependent on local heating temperature and analytical approach

The within-temperature associations between endothelium-dependent dilation and the NO-mediated component (Δ%max) during local heating to 39°C and 42°C are presented in Figure 3. Endothelium-dependent dilation was significantly related to NO-dependent dilation expressed as the difference between the local heating and L-NAME plateau (Δ%max) during 39°C (R^2^=0.79, P<0.001; Figure 3A) and 42°C (R^2^=0.56, P<0.001; Figure 3C). The within-temperature associations between endothelium-dependent dilation and the NO-mediated component for the additional analytical approaches to quantify NO-dependent dilation are shown in Supplemental Figure 1. Comparisons of between-temperature associations for endothelium-dependent dilation and each NO quantification are presented in Supplemental Table 3.

**Figure 3.**
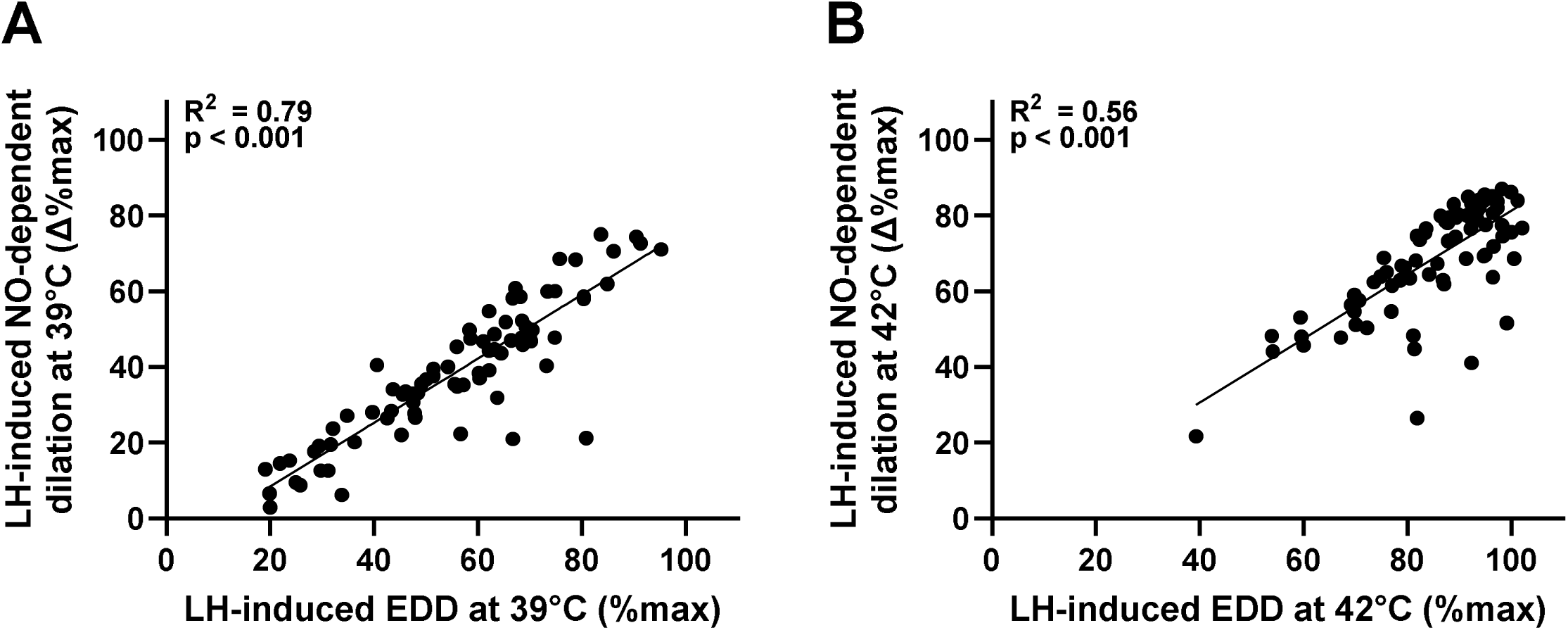
Relation between the magnitude of local heating (LH)-induced endothelium-dependent dilation (EDD) to the nitric oxide (NO)-mediated component (Δ%max) of that response at 39°C and 42°C. Associations presented within local heating protocol for 39°C (A) and 42°C (B). EDD was quantified as the local heat plateau (%max) and NO-dependent dilation was quantified as the difference between the local heat plateau and the L-NAME (N^G^-nitro-l-arginine methyl ester) plateau (Δ%max). Data were analyzed by simple linear regression.

### Acetylcholine-induced endothelium- and NO-dependent dilation

Endothelium-dependent dilation in response to acetylcholine in control and L-NAME-treated sites are shown in Figure 4A. NO synthase inhibition reduced the vasodilatory response to acetylcholine (main effect of site: P<0.001). NO-dependent dilation expressed as the difference in AUC (Δa.u.) between control and L-NAME sites is presented in Figure 4B. Fitted group and individually modeled dose-response curves are presented for each site in Figure 4C and 4D. Acetylcholine-induced NO-dependent dilation (Δ%NOpredicted) is presented in Figure 4E. NO-dependent dilation was lower at logEC_50_ compared with logEC_90_ (P<0.001). Parameters for fitted group curves are shown in Supplemental Table 4. Secondary quantifications of acetylcholine-induced NO-dependent dilation are presented in Supplemental Table 5.

**Figure 4.**
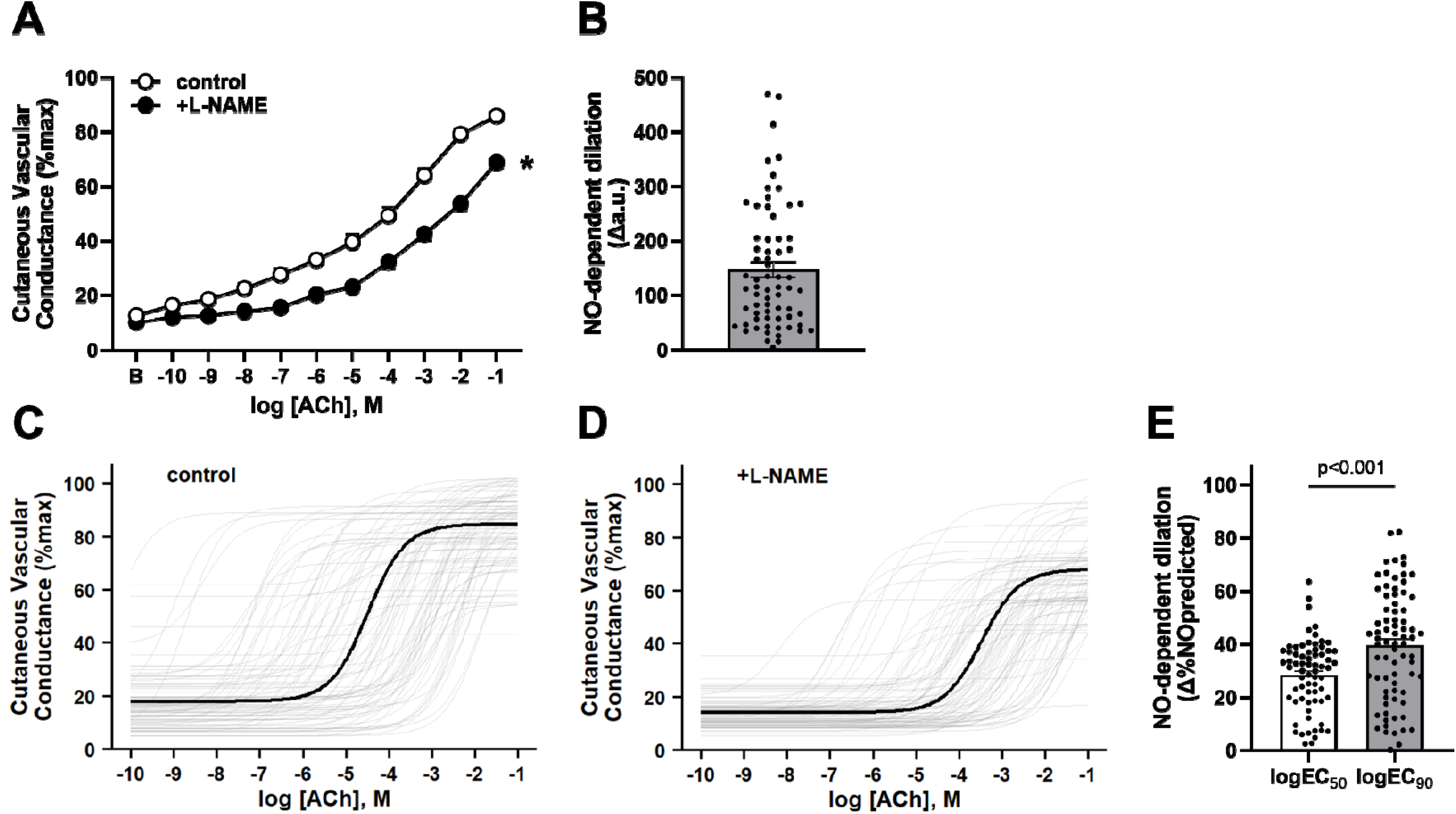
Assessments of acetylcholine-induced endothelium- and nitric oxide (NO)-dependent vasodilation in the cutaneous circulation. Endothelium-dependent dilation (cutaneous vascular conductance, %max) was quantified at each dose of acetylcholine at the control and NO-synthase (L-NAME; N^G^-nitro-l-arginine methyl ester) inhibited sites (A). NO-dependent dilation was calculated as the difference in area under the curve between control and L-NAME sites (B; Δa.u.). Fitted group and individually modeled dose-response curves are presented by control (C) and L-NAME (D) sites. NO-dependent dilation was calculated as the difference between control and L-NAME predicted responses (%max) at individual logEC_50_ and logEC_90_ (E; Δ%NO predicted). See *Analytical Procedures* for detailed mathematical approaches. Modeled curves parameters are listed in Supplemental Table 4. P-values determined by two-way repeated measures ANOVA (A) and paired *t*-test (E). *P<0.05 vs. control site. ACh, acetylcholine; a.u., arbitrary units.

### Microvascular endothelial responses to local heating and acetylcholine are not related within individuals

The linear associations between endothelium-dependent dilation in response to local heating and the perfusion of acetylcholine are shown in Table 1. Across all three approaches to quantify acetylcholine-induced endothelium-dependent dilation, none were related to local heating-induced endothelium-dependent dilation (all P>0.05). Table 2 presents correlation coefficients for NO-dependent dilation between local heating and acetylcholine stimuli. Regardless of how NO was quantified, there was no relation between local heating- or acetylcholine-induced responses (all P>0.05).

**Table 1.**
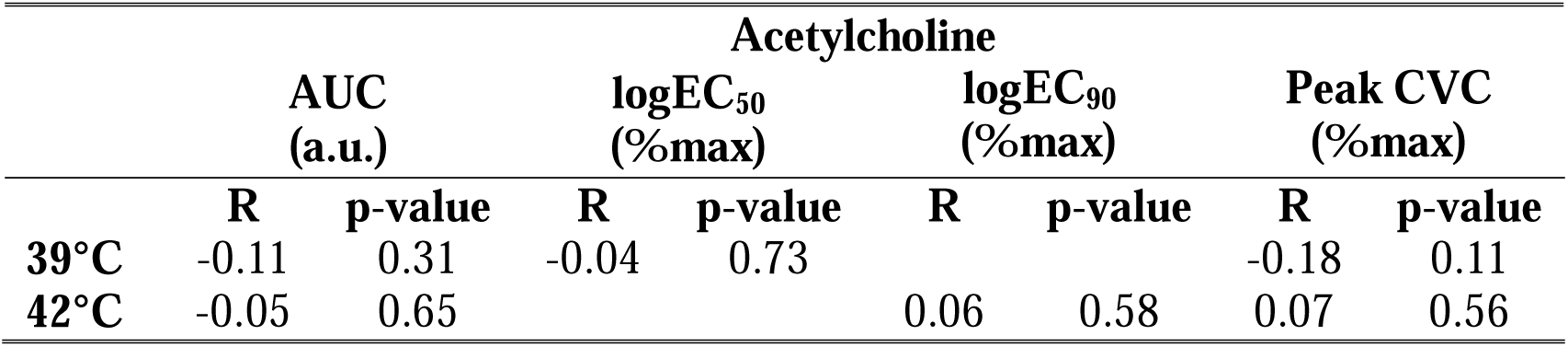
Correlation coefficients for the relation between endothelium-dependent dilation responses to acetylcholine and local heating to 39°C and 42°C (n=80). Data were analyzed by Pearson’s correlation. AUC, area under the curve; a.u., arbitrary units; CVC, cutaneous vascular conductance.

**Table 2.** Correlation coefficients for the relation between nitric oxide (NO)-dependent dilation responses to acetylcholine and local heating to 39°C and 42°C. Some data were excluded when the NO synthase inhibited site response exceeded the control site response, resulting in a negative NO-dependent dilation value. Final sample sizes for each comparison are presented in the table. Data were analyzed by Pearson’s correlation. AUC, area under the curve; a.u., arbitrary units; CVC, cutaneous vascular conductance.

|  |  | Acetylcholine |  |  |  |  |  |  |  |  |  |
| --- | --- | --- | --- | --- | --- | --- | --- | --- | --- | --- | --- |
| | | ΔAUC<br>(n=67) | | %AUC<br>(n=67) | | logEC<sub>50</sub><br>(n=69) | | logEC<sub>90</sub><br>(n=76) | | ΔPeak CVC<br>(n=72) | |
|  |  | R | p-value | R | p-value | R | p-value | R | p-value | R | p-value |
| 39°C | $\Delta\%$max | -0.09 | 0.49 | -0.17 | 0.18 | -0.07 | 0.57 | | | 0.13 | 0.29 |
|  | %plateau | 0.01 | 0.91 | -0.20 | 0.11 | -0.07 | 0.56 |  |  | -0.02 | 0.85 |
|  | %plateau<sub>BL</sub> | -0.03 | 0.82 | -0.15 | 0.23 | -0.04 | 0.76 |  |  | 0.01 | 0.91 |
| 42°C | $\Delta\%$max | 0.02 | 0.88 | -0.09 | 0.49 | | | 0.07 | 0.56 | 0.00 | 0.98 |
|  | %plateau | 0.20 | 0.11 | -0.07 | 0.59 |  |  | 0.20 | 0.08 | 0.11 | 0.36 |
|  | %plateau<sub>BL</sub> | 0.07 | 0.58 | -0.10 | 0.41 |  |  | 0.19 | 0.09 | 0.20 | 0.08 |

## DISCUSSION

The primary finding of this study is that cutaneous microvascular endothelial function measured in response to three commonly used stimuli are not comparable within an individual. Within local heating stimuli, we found that the magnitude of endothelium-dependent dilation was greater at 42°C compared to that at 39°C, and that the NO-dependent contribution to this response was also greater at 42°C, both when expressed as the difference between the local heating and L-NAME plateaus (Δ%max) and as percentage of the local heating plateau (%plateau). We further demonstrate that there is a significant relation between endothelium- and NO-dependent dilation (expressed as Δ%max) within each local heating temperature. Lastly, we found that endothelium- and NO-dependent dilation in response to either local heating protocol or exogenous acetylcholine perfusion were not related, regardless of how either response was quantified. Collectively, these data from a large sample size suggest that while all three techniques are valid for assessing microvascular endothelium- and NO-dependent dilation, the elicited responses are not comparable within an individual. Thus, careful consideration is warranted when directly comparing quantitative results from studies utilizing differing physiological and pharmacological stimuli to assess cutaneous vascular endothelial function.

Microvascular dysfunction and impaired endothelial signaling, including reduced NO bioavailability, are among the earliest manifestations of CVD and are potent predictors of CVD morbidity and mortality^1^. The human cutaneous circulation is commonly used to assess microvascular endothelial function and dysfunction in healthy individuals^26,32–34^, populations at high risk for CVD^3,29,35,36^, and in populations with overt clinical disease^6,9,23^. One commonly used assessment is local heating of the skin to 39°C or 42°C. Local heating produces a vasodilatory response that is reported to be 70-90% dependent on NO^15,16,31^. Recent reports suggest that the endothelium-dependent plateau in blood flow during local heating to 39°C induces a greater NO-dependent response than that at 42°C^14^, despite the overall magnitude of vasodilation being greater at 42°C. Our data similarly demonstrate that the magnitude of the endothelium-dependent plateau response is greater at 42°C compared with 39°C. In contrast, we show that the NO-dependent contribution to the local heating plateau was also greater during 42°C than 39°C when expressed as both Δ%max and %plateau. Further, we found no difference in NO-dependent dilation between temperatures when expressed as a percentage of the plateau above baseline (%plateau_BL_).

Local heating of the skin is a non-invasive, robust assessment of microvascular NO-dependent dilation in humans; however, direct quantification requires pharmacological NO synthase inhibition, often administered via intradermal microdialysis, which requires specialized training and resources. The local heating plateau at 39°C has recently been proposed as a non-invasive surrogate for NO-dependent dilation when direct quantification is not feasible^31^. In agreement, we found a strong relation between the endothelium-dependent plateau and the NO-dependent contribution of that response when expressed as Δ%max during local heating to both 39°C and 42°C. Similar to recent findings^16^, our data show a stronger association at 39°C compared with 42°C. Further, we found that NO-dependent dilation expressed as %plateau and %plateau_BL_ were associated with the endothelium-dependent plateau during local heating at 39°C but not at 42°C (Supplemental Figure 1). This supports recent data suggesting that the local heating plateau is more reflective of endothelium-derived NO at 39°C than at 42°C^14^.

Importantly, data from our larger sample extend the findings of Wong & Hayat^31^ by providing direct support for the local heating plateau at 39°C as a more reliable surrogate for NO-dependent dilation than the local heating plateau at 42°C when direct quantification with NO synthase inhibitors are not possible.

We next examined whether endothelial responses during local heating to 42°C were correlated with the responses at 39°C within individuals. Consistent with differences in the magnitude of the response, there was no relation between endothelium-dependent dilation between temperatures (Supplemental Table 3). Similarly, NO-dependent dilation expressed as Δ%max and %plateau were not significantly associated between temperatures. These findings suggest that responses to local heating to 42°C and 39°C are not related within an individual. Furthermore, given the lack of relation between stimuli, studies that utilize different local thermal stimuli are not interchangeable, and cross-study comparisons be made with caution.

Interestingly, we found a significant association between temperatures when NO-dependent dilation was expressed as %plateau_BL_. This finding, however, likely reflects a limitation of the mathematical approach rather than a true association. Quantification of NO-dependent dilation as %plateauBL was more likely to approach or reach 100% when the L-NAME plateau was close to or lower than baseline, resulting in a clustering of values at 100% for both temperatures (Figure 2C). This approach thus overestimates NO-dependent dilation, consistent with findings reported by others^16,31^. Collectively, these data suggest that local heating to 42°C produces greater endothelium- and NO-dependent dilation than that at 39°C, and that responses to 42°C do not predict those at 39°C within individuals. Given that findings and interpretations may differ based on the analytical approach, we refer the reader to the analysis by Wolf et al. for specific recommendations^16^.

Of note, relative CVC responses at the local heating plateau at 39°C were normally distributed across participants, ranging from 19-95%max, indicating a substantial amount of between-person variability in endothelium-dependent dilation. Although the 42°C local heating plateau values spanned a similar range (39-100%max), 75% (59/80) fell within one standard deviation of the mean (85±13%max), suggesting that 42°C elicits a more uniform endothelium-dependent response in young healthy adults. Collectively, these findings suggest that local heating to 42°C is a more robust and consistent stimulus for the assessment of microvascular endothelial function, particularly when NO synthase inhibitors can be employed to directly quantify NO-dependent dilation. However, when it is not feasible to couple local heating with intradermal microdialysis perfusion of NO synthase inhibitors, the local heating plateau at 39°C serves as a more appropriate surrogate for estimating NO-dependent dilation, despite having greater variability in quantified endothelium-dependent dilation.

Perfusion of exogenous acetylcholine induces endothelium-dependent vasodilation that is mediated by NO, prostaglandins, and other endothelial-derived relaxing factors^37,38^. Although the NO-dependent contribution to endothelium-dependent dilation during acetylcholine perfusion (∼30-40%) is less than that with local heating (∼70-90%), exogenous acetylcholine is widely used in *in vitro* and preclinical studies; thus, its application in the human cutaneous microvasculature allows for direct cross-model comparison and translation of mechanistic studies of endothelial function. However, quantifying NO-dependent responses to exogenous acetylcholine is difficult, as the contribution of NO varies depending on the duration and dose employed^24,39^, and no standardized approach exists for calculating and interpreting these responses. NO-dependent dilation in response to acetylcholine is commonly quantified as the difference between control and L-NAME sites based on calculated AUC^5,21,29,40^, the peak vasodilatory response^17,41^, or differences in modeling parameters such as logEC_50_ or Hill Slope^17,29,42^, all of which allow for between group and/or treatment comparisons based on the study design. In the current study, we sought to leverage our larger sample size to present values for commonly reported calculations of NO-dependent dilation, and, further, to evaluate a novel approach based on pharmacological curve fitting. Given that reported NO-dependent dilation is heavily dependent on the mathematical approach employed (Supplemental Table 5), the quantification of acetylcholine-induced NO-dependent dilation must be chosen based on the study design and research question at hand.

Studies utilizing both local heating and acetylcholine dose-responses concurrently have found that microvascular responses to these stimuli agree when comparing between groups. For example, responses to both acetylcholine and local heating are reduced in the cutaneous microvasculature of healthy older compared to young adults^19,20,37,43,44^. Furthermore, we have found reductions in endothelium- and NO-dependent dilation to both local heating to 42°C and acetylcholine in women with a history of preeclampsia, and that responses to both stimuli are similarly sensitive to mechanistic intervention in these women^3,17,21,41,42,45,46^. However, no studies to date have examined if microvascular endothelial responses to these stimuli are directly comparable within an individual. Here, we are the first to demonstrate that microvascular endothelium- and NO-dependent dilation responses to local heating and exogenous acetylcholine are not comparable within an individual. Moreover, our data show that regardless of how NO-dependent dilation was quantified, no relation exists between local heating- and acetylcholine-induced dilation. Given that the relative contribution of NO to local heating- and acetylcholine-induced dilation are vastly different, and that these stimuli rely on different intracellular signaling mechanisms to induce vasodilation, it is perhaps not surprising that NO-dependent dilation between stimuli are not related. To allow for a more meaningful comparison, we additionally quantified acetylcholine endothelium- and NO-dependent dilation at each individual’s logEC_50_ and logEC_90_, which we hypothesized may be more representative of the local heating-induced vasodilatory responses at 39°C and 42°C, respectively. However, despite employing advanced analytical approaches to standardize NO-dependent dilation responses, we found no relation between acetylcholine- and local heating-mediated endothelium- or NO-dependent dilation. Therefore, our findings suggest that while all three stimuli assess endothelium- and NO-dependent dilation in the microvasculature, direct comparison of the magnitude of the responses across these commonly used techniques should be performed with caution.

### Perspectives

Several studies over recent decades have utilized the cutaneous microvasculature as a representative in vivo model to examine endothelium- and NO-dependent dilation in humans. These studies have consistently found reduced microvascular endothelial function in populations with high CVD risk or overt disease. Three endothelium-dependent stimuli have predominated the literature, yet it remains unclear whether these stimuli are reflective of each other, limiting cross-study comparisons. Therefore, the overarching goal of this study was to directly examine the relation between commonly used endothelium-dependent stimuli in the cutaneous microvasculature. Our data demonstrate that common physiological (local heating) and pharmacological (acetylcholine) stimuli for assessing cutaneous microvascular endothelium- and NO-dependent dilation are not comparable within an individual, and cross-study comparisons using different stimuli should be avoided. Although local heating to 39°C and 42°C and perfusion of acetylcholine all induce robust endothelium-dependent dilation in the cutaneous circulation, each has a differing NO-dependent contribution to the overall vasodilation response. Thus, the endothelium-dependent stimuli and mathematical approaches utilized should be guided by the research question of interest.

## Data Availability

All data produced in the present study are available upon reasonable request to the authors

## Non-standard Abbreviations and Acronyms

AUC: area under the curve
a.u.: arbitrary units
BMI: body mass index
CVC: cutaneous vascular conductance
CVD: cardiovascular disease
L-NAME: N^G^-nitro-L-arginine methyl ester
LH: local heating
MAP: mean arterial pressure
NO: nitric oxide

## Author Contributions

**KSS**: acquisition of data, analysis and interpretation of data, conception and design, manuscript preparation; **MGE**: acquisition of data, analysis and interpretation of data, manuscript preparation; **CEG**: acquisition of data, manuscript preparation; **JG**: analysis and interpretation of data, conception and design, manuscript preparation; **AES**: analysis and interpretation of data, conception and design, manuscript preparation

## Acknowledgments

The authors acknowledge Aaron Autler, Ruda Lee, Grace Maurer, Joy Mochache, and Navya Vadlamudi for their assistance with data collection and the participants for contributing their time and effort to the completion of this project.

## REFERENCES

1. Holowatz LA, Thompson-Torgerson CS, Kenney WL. The human cutaneous circulation as a model of generalized microvascular function. J Appl Physiol (1985). 2008;105:370–372. doi: 10.1152/japplphysiol.00858.2007

2. Debbabi H, Bonnin P, Ducluzeau PH, Lefthériotis G, Levy BI. Noninvasive assessment of endothelial function in the skin microcirculation. Am J Hypertens. 2010;23:541–546. doi: 10.1038/ajh.2010.10

3. Schwartz KS, Sun M, Jalal DI, Santillan MK, Stanhewicz AE. Reduced AT(2)R Signaling Contributes to Endothelial Dysfunction After Preeclampsia. Hypertension. 2025;82:904–913. doi: 10.1161/hypertensionaha.124.24098

4. Greaney JL, Stanhewicz AE, Kenney WL. Chronic statin therapy is associated with enhanced cutaneous vascular responsiveness to sympathetic outflow during passive heat stress. J Physiol. 2019;597:4743–4755. doi: 10.1113/jp278237

5. Schwartz KS, Hernandez PV, Maurer GS, Wetzel EM, Sun M, Jalal DI, Stanhewicz AE. Impaired microvascular insulin-dependent dilation in women with a history of gestational diabetes. Am J Physiol Heart Circ Physiol. 2024;327:H793–h803. doi: 10.1152/ajpheart.00223.2024

6. DuPont JJ, Ramick MG, Farquhar WB, Townsend RR, Edwards DG. NADPH oxidase-derived reactive oxygen species contribute to impaired cutaneous microvascular function in chronic kidney disease. Am J Physiol Renal Physiol. 2014;306:F1499–1506. doi: 10.1152/ajprenal.00058.2014

7. Green DJ, Maiorana AJ, Siong JH, Burke V, Erickson M, Minson CT, Bilsborough W, O’Driscoll G. Impaired skin blood flow response to environmental heating in chronic heart failure. Eur Heart J. 2006;27:338–343. doi: 10.1093/eurheartj/ehi655

8. Andersson SE, Edvinsson ML, Edvinsson L. Cutaneous vascular reactivity is reduced in aging and in heart failure: association with inflammation. Clin Sci (Lond). 2003;105:699–707. doi: 10.1042/cs20030037

9. Greaney JL, Kutz JL, Shank SW, Jandu S, Santhanam L, Alexander LM. Impaired Hydrogen Sulfide-Mediated Vasodilation Contributes to Microvascular Endothelial Dysfunction in Hypertensive Adults. Hypertension. 2017;69:902–909. doi: 10.1161/hypertensionaha.116.08964

10. Landmesser U, Hornig B, Drexler H. Endothelial function: a critical determinant in atherosclerosis? Circulation. 2004;109:Ii27-33. doi: 10.1161/01.CIR.0000129501.88485.1f

11. Sitia S, Tomasoni L, Atzeni F, Ambrosio G, Cordiano C, Catapano A, Tramontana S, Perticone F, Naccarato P, Camici P, et al. From endothelial dysfunction to atherosclerosis. Autoimmun Rev. 2010;9:830–834. doi: 10.1016/j.autrev.2010.07.016

12. Ashor AW, Lara J, Siervo M, Celis-Morales C, Oggioni C, Jakovljevic DG, Mathers JC. Exercise modalities and endothelial function: a systematic review and dose-response meta-analysis of randomized controlled trials. Sports Med. 2015;45:279–296. doi: 10.1007/s40279-014-0272-9

13. Daiber A, Steven S, Weber A, Shuvaev VV, Muzykantov VR, Laher I, Li H, Lamas S, Münzel T. Targeting vascular (endothelial) dysfunction. Br J Pharmacol. 2017;174:1591–1619. doi: 10.1111/bph.13517

14. Choi PJ, Brunt VE, Fujii N, Minson CT. New approach to measure cutaneous microvascular function: an improved test of NO-mediated vasodilation by thermal hyperemia. J Appl Physiol (1985). 2014;117:277–283. doi: 10.1152/japplphysiol.01397.2013

15. Minson CT, Berry LT, Joyner MJ. Nitric oxide and neurally mediated regulation of skin blood flow during local heating. J Appl Physiol (1985). 2001;91:1619–1626. doi: 10.1152/jappl.2001.91.4.1619

16. Wolf ST, Dillon GA, Alexander LM, Kenney WL, Stanhewicz AE. Quantification and interpretation of nitric oxide-dependent cutaneous vasodilation during local heating. J Appl Physiol (1985). 2024;137:1418–1424. doi: 10.1152/japplphysiol.00558.2024

17. Stanhewicz AE, Jandu S, Santhanam L, Alexander LM. Increased Angiotensin II Sensitivity Contributes to Microvascular Dysfunction in Women Who Have Had Preeclampsia. Hypertension. 2017;70:382–389. doi: 10.1161/hypertensionaha.117.09386

18. Medow MS, Glover JL, Stewart JM. Nitric oxide and prostaglandin inhibition during acetylcholine-mediated cutaneous vasodilation in humans. Microcirculation. 2008;15:569–579. doi: 10.1080/10739680802091526

19. Minson CT, Holowatz LA, Wong BJ, Kenney WL, Wilkins BW. Decreased nitric oxide-and axon reflex-mediated cutaneous vasodilation with age during local heating. J Appl Physiol (1985). 2002;93:1644–1649. doi: 10.1152/japplphysiol.00229.2002

20. Wenner MM, Sebzda KN, Kuczmarski AV, Pohlig RT, Edwards DG. ET(B) receptor contribution to vascular dysfunction in postmenopausal women. Am J Physiol Regul Integr Comp Physiol. 2017;313:R51–r57. doi: 10.1152/ajpregu.00410.2016

21. Schwartz KS, Jalal DI, Stanhewicz AE. Oral losartan treatment improves microvascular endothelial function via nitric oxide-dependent mechanisms in women with a history of preeclampsia. Am J Hypertens. 2025. doi: 10.1093/ajh/hpaf033

22. Dillon GA, Stanhewicz AE, Serviente C, Flores VA, Stachenfeld N, Alexander LM. Seven days of statin treatment improves nitric-oxide mediated endothelial-dependent cutaneous microvascular function in women with endometriosis. Microvasc Res. 2022;144:104421. doi: 10.1016/j.mvr.2022.104421

23. Smith CJ, Santhanam L, Bruning RS, Stanhewicz A, Berkowitz DE, Holowatz LA. Upregulation of inducible nitric oxide synthase contributes to attenuated cutaneous vasodilation in essential hypertensive humans. Hypertension. 2011;58:935–942. doi: 10.1161/hypertensionaha.111.178129

24. Craighead DH, Smith CJ, Alexander LM. Blood pressure normalization via pharmacotherapy improves cutaneous microvascular function through NO-dependent and NO-independent mechanisms. Microcirculation. 2017;24. doi: 10.1111/micc.12382

25. Stewart JM, Nafday A, Ocon AJ, Terilli C, Medow MS. Cutaneous constitutive nitric oxide synthase activation in postural tachycardia syndrome with splanchnic hyperemia. Am J Physiol Heart Circ Physiol. 2011;301:H704–711. doi: 10.1152/ajpheart.00171.2011

26. Stanhewicz AE, Greaney JL, Kenney WL, Alexander LM. Sex- and limb-specific differences in the nitric oxide-dependent cutaneous vasodilation in response to local heating. Am J Physiol Regul Integr Comp Physiol. 2014;307:R914–919. doi: 10.1152/ajpregu.00269.2014

27. Greaney JL, DuPont JJ, Lennon-Edwards SL, Sanders PW, Edwards DG, Farquhar WB. Dietary sodium loading impairs microvascular function independent of blood pressure in humans: role of oxidative stress. J Physiol. 2012;590:5519–5528. doi: 10.1113/jphysiol.2012.236992

28. Greaney JL, Saunders EFH, Alexander LM. Short-term salicylate treatment improves microvascular endothelium-dependent dilation in young adults with major depressive disorder. Am J Physiol Heart Circ Physiol. 2022;322:H880–h889. doi: 10.1152/ajpheart.00643.2021

29. Greaney JL, Saunders EFH, Santhanam L, Alexander LM. Oxidative Stress Contributes to Microvascular Endothelial Dysfunction in Men and Women With Major Depressive Disorder. Circ Res. 2019;124:564–574. doi: 10.1161/circresaha.118.313764

30. Turner CG, Hayat MJ, Grosch C, Quyyumi AA, Otis JS, Wong BJ. Endothelin A receptor inhibition increases nitric oxide-dependent vasodilation independent of superoxide in non-Hispanic Black young adults. J Appl Physiol (1985). 2023;134:891–899. doi: 10.1152/japplphysiol.00739.2022

31. Wong BJ, Hayat MJ. Reliability of using the skin blood flow plateau response to local heating as a noninvasive assessment of nitric oxide-dependent vasodilation. J Appl Physiol (1985). 2025;139:454–464. doi: 10.1152/japplphysiol.00324.2025

32. Schwartz KS, Lang JA, Stanhewicz AE. Angiotensin II type 2 receptor-mediated dilation is greater in the cutaneous microvasculature of premenopausal women compared with men. J Appl Physiol (1985). 2023;135:1236–1242. doi: 10.1152/japplphysiol.00382.2023

33. Kellogg DL, Jr., Zhao JL, Wu Y. Roles of nitric oxide synthase isoforms in cutaneous vasodilation induced by local warming of the skin and whole body heat stress in humans. J Appl Physiol (1985). 2009;107:1438–1444. doi: 10.1152/japplphysiol.00690.2009

34. Greaney JL, Surachman A, Saunders EFH, Alexander LM, Almeida DM. Greater Daily Psychosocial Stress Exposure is Associated With Increased Norepinephrine-Induced Vasoconstriction in Young Adults. J Am Heart Assoc. 2020;9:e015697. doi: 10.1161/jaha.119.015697

35. Schwartz K, Campbell N, Jalal D, Stanhewicz A. Reductions in angiotensin II type 2 receptor-mediated vasodilation contribute to increased angiotensin II vasoconstrictor sensitivity in women with preeclampsia history. Clinical Science. 2025. doi: 10.1042/cs20245238

36. Halstead KM, Wetzel EM, Cho JL, Stanhewicz AE. Sex Differences in Oxidative Stress-Mediated Reductions in Microvascular Endothelial Function in Young Adult e-Cigarette Users. Hypertension. 2023;80:2641–2649. doi: 10.1161/hypertensionaha.123.21684

37. Holowatz LA, Thompson CS, Minson CT, Kenney WL. Mechanisms of acetylcholine-mediated vasodilatation in young and aged human skin. J Physiol. 2005;563:965–973. doi: 10.1113/jphysiol.2004.080952

38. Kellogg DL, Jr., Zhao JL, Coey U, Green JV. Acetylcholine-induced vasodilation is mediated by nitric oxide and prostaglandins in human skin. J Appl Physiol (1985). 2005;98:629–632. doi: 10.1152/japplphysiol.00728.2004

39. Brunt VE, Fujii N, Minson CT. Endothelial-derived hyperpolarization contributes to acetylcholine-mediated vasodilation in human skin in a dose-dependent manner. J Appl Physiol (1985). 2015;119:1015–1022. doi: 10.1152/japplphysiol.00201.2015

40. Dillon GA, Greaney JL, Shank S, Leuenberger UA, Alexander LM. AHA/ACC-defined stage 1 hypertensive adults do not display cutaneous microvascular endothelial dysfunction. Am J Physiol Heart Circ Physiol. 2020;319:H539–h546. doi: 10.1152/ajpheart.00179.2020

41. Stanhewicz AE, Dillon GA, Serviente C, Alexander LM. Acute systemic inhibition of inflammation augments endothelium-dependent dilation in women with a history of preeclamptic pregnancy. Pregnancy Hypertens. 2022;27:81–86. doi: 10.1016/j.preghy.2021.12.010

42. Lee R, Greaney JL, Santillan MK, Pierce GL, Stanhewicz AE. Vascular endothelial function is preserved in women with a history of preeclampsia currently receiving antidepressant pharmacotherapy. Am J Physiol Heart Circ Physiol. 2026;330:H524–h530. doi: 10.1152/ajpheart.00890.2025

43. Mitchell UH, Burton S, Gordon C, Mack GW. The Effect of Being Aerobically Active vs. Inactive on Cutaneous Vascular Conductance during Local Heat Stress in an Older Population. Front Physiol. 2017;8:859. doi: 10.3389/fphys.2017.00859

44. Serviente C, Berry CW, Kenney WL, Alexander LM. Healthy active older adults have enhanced K(+) channel-dependent endothelial vasodilatory mechanisms. Am J Physiol Regul Integr Comp Physiol. 2020;319:R19–r25. doi: 10.1152/ajpregu.00049.2020

45. Stanhewicz AE, Jandu S, Santhanam L, Alexander LM. Alterations in endothelin type B receptor contribute to microvascular dysfunction in women who have had preeclampsia. Clin Sci (Lond). 2017;131:2777–2789. doi: 10.1042/cs20171292

46. Stanhewicz AE, Alexander LM. Local angiotensin-(1-7) administration improves microvascular endothelial function in women who have had preeclampsia. Am J Physiol Regul Integr Comp Physiol. 2020;318:R148–r155. doi: 10.1152/ajpregu.00221.2019

47. Ketel IJ, Stehouwer CD, Serné EH, Poel DM, Groot L, Kager C, Hompes PG, Homburg R, Twisk JW, Smulders YM, et al. Microvascular function has no menstrual-cycle-dependent variation in healthy ovulatory women. Microcirculation. 2009;16:714–724. doi: 10.3109/10739680903199186

48. D’Agata MN, Hoopes EK, Berube FR, Hirt AE, Kuczmarski AV, Ranadive SM, Wenner MM, Witman MA. Evidence of reduced peripheral microvascular function in young Black women across the menstrual cycle. J Appl Physiol (1985). 2021;131:1783–1791. doi: 10.1152/japplphysiol.00452.2021

49. Williams JS, Dunford EC, MacDonald MJ. Impact of the menstrual cycle on peripheral vascular function in premenopausal women: systematic review and meta-analysis. Am J Physiol Heart Circ Physiol. 2020;319:H1327–h1337. doi: 10.1152/ajpheart.00341.2020

50. Wolf ST, Jablonski NG, Ferguson SB, Alexander LM, Kenney WL. Four weeks of vitamin D supplementation improves nitric oxide-mediated microvascular function in college-aged African Americans. Am J Physiol Heart Circ Physiol. 2020;319:H906–h914. doi: 10.1152/ajpheart.00631.2020

51. Wenner MM, Wilson TE, Davis SL, Stachenfeld NS. Pharmacological curve fitting to analyze cutaneous adrenergic responses. J Appl Physiol (1985). 2011;111:1703–1709. doi: 10.1152/japplphysiol.00780.2011

