## Supplemental Material for "Microvascular Responses to Common Endothelial Stimuli Are Not Related in Humans"

**Detailed Methods**

**Ethical Approval**

The data that support the findings of this study are available from the corresponding author upon reasonable request. Written and verbal informed consent were obtained prior to study enrollment in accordance with the World Medical Association Declaration of Helsinki. All experimental protocols were approved by the University of Iowa Institutional Review Board (IRB; no. 202309383) and the University of Delaware IRB (no. 2190901). The use of pharmacological agents with intradermal microdialysis was approved by the U.S. Food and Drug Administration (IND no. 156,345).

**Participants**

A total of 80 young adults (40 males/40 females) completed the study, with 40 participants (20 males/20 females) tested at each study site. Participants were recruited by public advertisement in Iowa City, IA and Newark, DE. All participants were 18-30 years of age and completed a medical screening that included a physical examination, a collection of health history, and a urine pregnancy test (females). Exclusion criteria included current or past cardiovascular, renal, and/or metabolic disease, current use of anti-hypertensive or cholesterol-lowering medications, and/or current pregnancy. Race and ethnicity were self-reported by all participants.

**Microvascular Reactivity Measures**

Participants completed one experimental visit where they were asked to fast and withhold caffeine for 12 hours prior and refrain from strenuous physical activity and alcohol for 24 hours prior. Given that microvascular endothelial function is not influenced by menstrual cycle phase^47-49^, females were tested without regard to menstruation.

Participants were instrumented in a semi-recumbent position and remained in this position for the duration of the visit. Following 5 minutes of local ice application to anesthetize the skin, four intradermal microdialysis fibers (CMA 31 Linear Microdialysis Probe, CMA Microdialysis, Holliston, MA) separated by ≥4cm were aseptically placed in the left ventral forearm for the local delivery of pharmacological agents. Pharmacological agents were weighed just prior to use, mixed with lactated Ringer’s solution, sterilized with syringe microfilters (Acrodisc; Pall, Ann Arbor, MI), wrapped in foil to prevent photodegradation, and perfused through each fiber at a rate of 2 μL/min (Bee Hive controller and Baby Bee microinfusion pumps; Bioanalytical Systems, West Lafayette, IN). Laser-Doppler flowmetry probes were placed in local heaters (Moor Instruments, Wilmington, DE) set to thermoneutral (33°C) directly over each microdialysis site to continuously measure cutaneous red cell flux within the tissue (~1mm3) treated by the microdialysis perfusates. Automated brachial blood pressure and heart rate (SureSigns VS2+, Philips Healthcare, Andover, MA and Connex Spot Monitor, Welch Allyn, Skaneateles Falls, NY) were measured every 4-5 minutes throughout the protocol. Blood pressure was measured in the contralateral arm at heart level. Following an initial hyperemia-resolution period (~60 minutes), baseline measurements were collected (~10 minutes), and endothelium- and NO-dependent dilation were subsequently assessed as described below.

*Physiological endothelium-dependent responses induced by local heating.* Two microdialysis fibers were randomly selected to be perfused with lactated Ringer’s (control) to undergo local heating to either 39°C or 42°C. Following baseline measurements, the temperature of the local heaters over each site were raised to the appropriate temperature following a standardized rapid local heating protocol (0.1°C·s^-1^)^3,16,26,27^ and remained at the site-specific temperature until maximal skin blood flow (below) was performed. Following the plateau in skin blood flow associated with local heating (~40 minutes), both sites were then perfused with 15 mM N^G^-nitro-L-arginine methyl ester (L-NAME; Calbiochem, EMD Millipore, Billerica, MA) at 4 μL/min until a second plateau in skin blood flow was reached (~40 minutes) to determine site-specific NO-dependent dilation. Maximal skin blood flow commenced following completion of the local heating protocols.

*Pharmacological endothelium-dependent dilation induced by acetylcholine.* The remaining two microdialysis fibers were randomly selected to be perfused with either lactated Ringer’s or 15 mM L-NAME. Following baseline measurements, ascending concentrations of acetylcholine (10^-10^-10^-1^ M; United States Pharmacopeia, Rockville, MD) alone (control) or in combination with L-NAME were perfused sequentially for 5 minutes each^5,21,28,29^. Maximal skin blood flow commenced following completion of the dose-response protocol.

*Maximal skin blood flow.* Following completion of the physiological and pharmacological protocols, all microdialysis fibers were perfused with 28 mM sodium nitroprusside (United States Pharmacopeia) at 4 μL/min and all local heaters were simultaneously increased to 43°C until a plateau associated with maximal blood flow was obtained at each site (~20 minutes)^3,14,16,19,20,26,27^.

**Analytical Procedures**

All data collection and analysis procedures were standardized prior to testing. Data were recorded at 40 Hz and stored for offline analysis (PowerLab and LabChart; AD Instruments, Sydney, Australia). Absolute cutaneous vascular conductance was calculated (CVC=laser-Doppler flux/mean arterial pressure) and normalized to a percentage of site-specific maximum (relative CVC, %max) for analysis^3,5,9,14-21,26-30^.

*Quantification of local heating-induced responses:* Local heating (LH)-induced endothelium-dependent dilation was quantified during a stable 3-5 min plateau in skin blood flow during local heating^3,26,27^. The primary quantification of NO-dependent dilation was calculated at each site as the difference between the local heating- and the L-NAME-induced plateaus in skin blood flow [NO {∆%max} = LH plateau – L-NAME plateau]. This quantification is most commonly used to report the NO-dependent component of the local heating response^3,16,17,26,27,31,50^. To enhance rigor and transparency, local heating-induced NO-dependent dilation was also calculated as (1) a percentage of the local heating plateau [NO {%plateau} = (LH plateau – L-NAME plateau) / (LH plateau) x 100] and (2) a percentage of the local heating plateau above baseline [NO {%plateau_BL_} = [(LH plateau − baseline) − (L-NAME plateau − baseline)] / (LH plateau − baseline) × 100], according to recent recommendations^16,31^.

*Pharmacological Curve Modeling:* Acetylcholine-induced endothelium-dependent dilation was assessed during a stable 1-2-min plateau in skin blood flow for each concentration of acetylcholine^5,21,28,42^. Acetylcholine dose-response curves were fitted for the entire sample using a four-parameter nonlinear mixed effects model with Hill slope constrained to 1^51^. Group-level parameters were estimated (fixed effects: top, bottom, logEC_50_) and individual curves were fit based on the deviation from the fitted group curve parameters (random effects: top, bottom, logEC_50_). This analysis allowed for individual dose-response curves to be fitted for all participants (n=80) for both the acetylcholine alone and acetylcholine+L-NAME sites. Because this approach simultaneously estimates both group and individual parameters, statistical power is improved and potential bias resulting from having to exclude data from individuals whose data cannot be fit using the traditional two-step modeling approach is reduced^51^. Individual parameters logEC_50_ and logEC_90_ were calculated from the control site dose-response curves for each participant and were used for all subsequent analyses. The parameters logEC_50_ and logEC_90_ were selected given the approximate similarity between CVC (%max) at these estimated parameters and the local heating-induced endothelium-dependent plateaus in CVC (%max) at 39°C and 42°C for the full sample, respectively (see Figure 1). Then, using the individual logEC_50_ and logEC_90_ doses derived from the control site dose-response curves, the predicted relative CVC response (%max) was extracted from both the control and L-NAME dose-response curves to allow for the calculation of NO-dependent dilation. In relevant analyses described below, these predicted CVC values (%max) at the logEC_50_ and the logEC_90_ for each individual were compared to the CVC values (%max) in response to local heating to 39°C and 42°C, respectively.

*Quantification of acetylcholine-induced responses:* Given that there is no standardized approach for calculating acetylcholine-induced endothelial responses in the cutaneous vasculature, we quantified acetylcholine-induced endothelium- and NO-dependent dilation by methods consistently reported in the literature^5,9,17,21,42^. Individual acetylcholine-induced endothelium-dependent dilation was quantified from the control site dose-response data in several ways: (1) as the area under the curve (AUC; arbitrary units, a.u.) while accounting for baseline blood flow (GraphPad Prism 10.6.0, San Diego, CA); (2) the predicted CVC (%max) at individual logEC_50_ and logEC_90_ (see Pharmacological Curve Modeling above); and (3) the peak CVC (%max), defined as the maximum vasodilatory response, regardless of the dose at which it occurred. For each of these analytical approaches, we then quantified acetylcholine-induced NO-dependent dilation as: (1) the difference in AUC between sites [NO {∆a.u.} = control AUC – L-NAME AUC]; (2) the difference between the predicted CVC (%max) at the logEC_50_ or logEC_90_ between sites [NO {∆%NOpredicted} = predicted control CVC – predicted L-NAME CVC at the same dose]; (3) the difference in AUC between sites calculated as a percentage of the control site AUC [NO {%AUC} = (control AUC – L-NAME AUC) / (control AUC) x 100]; and (4) the difference between sites at the acetylcholine dose that elicited the peak CVC response at the control site [NO {∆%max} = peak control CVC – L-NAME CVC at the same dose]. Acetylcholine-induced NO-dependent dilation quantified as the difference in AUC (∆a.u) and the difference in predicted CVC (%NOpredicted) based on pharmacological modeling were employed as primary analyses. The additional quantification approaches were included to provide a more thorough analysis. We were unable to quantify NO-dependent dilation in all participants in instances when relative CVC (%max) responses at the L-NAME site were greater than those at the control site. As such, final sample sizes used for each NO quantification are presented in Supplemental Table 4.

All analyses were performed using R (version 4.5.2, R Foundation for Statistical Computing, Vienna, Austria; packages: dplyr, lme4, lmer, emmeans, ggplot2) unless otherwise noted. All data were assessed for normality via visual inspection of Q-Q plots, and sensitivity analysis confirmed results were robust to violations of homoscedasticity. One-way repeated measures ANOVAs were used to assess differences between local heating protocols (39°C vs. 42°C) for (1) local heating phases (baseline, peak, local heat plateau, L-NAME plateau) and (2) NO-dependent dilation calculated using each mathematical approach. A two-way repeated measures ANOVA was used to detect differences between acetylcholine dose-response curves (control vs. L-NAME). A paired t-test was used to compare NO-dependent dilation expressed as %NOpredicted between logEC_50_ and logEC_90_. Simple linear regression analyses were used to evaluate the associations between endothelium- and NO-dependent dilation between 39°C and 42°C and within each physiological and pharmacological protocol. Pearson’s correlation analyses were used to assess relations between vascular responses to local heating and acetylcholine. When significant main effects were identified, post hoc Tukey corrections were applied for specific planned comparisons as appropriate. Data are expressed as mean ± SD in tables and mean ± SEM in figures unless otherwise noted. Individual values are presented within each figure when appropriate.

| Participants, *n* | 80 |
| --- | --- |
| Age, y | 22±3 |
| Race, *n* (%) |  |
| White | 59 (74) |
| Black | 5 (6) |
| Asian | 12 (15) |
| >1 Race | 3 (4) |
| Unknown/not reported | 1 (1) |
| Ethnicity, *n* (%) |  |
| Not Hispanic or Latino | 69 (86) |
| Hispanic or Latino | 10 (13) |
| Unknown/not reported | 1 (1) |
| BMI, kg/m^2^ | 24.7±3.5 |
| Height (cm) | 171.9±10.2 |
| Weight (kg) | 73.2±14.4 |
| Resting mean arterial pressure, mmHg | 85±9 |
| Systolic | 116±12 |
| Diastolic | 71±10 |
| Resting heart rate, bpm | 70±12 |

**Supplemental Table 1. Participant demographics and resting hemodynamics.** BMI, body mass index.

| **Microdialysis Site** |  |
| --- | --- |
| **Acetylcholine Dose Response** |  |
| Lactated Ringer’s |  |
| baseline | 0.3±0.2 |
| maximum | 2.7±1.4 |
| L-NAME |  |
| baseline | 0.2±0.1 |
| maximum | 2.5±1.2 |
| **Local Heating** |  |
| 39°C |  |
| baseline | 0.3±0.3 |
| maximum | 2.2±1.2 |
| 42°C |  |
| baseline | 0.2±0.2 |
| maximum | 2.2±1.1 |

**Supplemental Table 2. Baseline and maximal absolute cutaneous vascular conductance (flux·mmHg^-1^).** Cutaneous Vascular Conductance (CVC)=laser-Doppler flux/mean arterial pressure. L-NAME, N^G^-nitro-l-arginine methyl ester. There were no differences between sites (all P>0.05).

|  |  |  | **39°C** | |
| --- | --- | --- | --- | --- |
|  |  | **Measurement** | **R^2^** | **p-value** |
| **42°C** | **EDD** | **local heat plateau** | 0.003 | 0.63 |
|  | **NO** | **∆%max** | 0.010 | 0.39 |
|  |  | **%plateau** | 0.039 | 0.08 |
|  |  | **%plateau_BL_** | 0.143 | **<0.01** |

**Supplemental Table 3. Associations between endothelium-dependent dilation (EDD) and nitric oxide (NO)-dependent dilation during local heating to 39°C and 42°C (n=80).** See *Analytical Procedures* for full information of how EDD and each index of NO was calculated. Data were analyzed by simple linear regression.

|  | **Control** | **+L-NAME** |
| --- | --- | --- |
| **Minimum, bottom** | 17.9 ± 1.3 [15.3 to 20.5] | 14.1 ± 0.7 [12.6 to 15.5] |
| **Maximum, top** | 84.5 ± 1.7 [81.1 to 87.9] | 67.9 ± 2.1 [63.8 to 71.9] |
| **Hill Slope** | 1.0 | 1.0 |
| **LogEC_50_** | -4.5 ± 0.2 [-4.9 to -4.1] | -3.5 ± 0.2 [-3.8 to -3.1] |
| **LogEC_90_** | -3.5 ± 0.2 [-4.0 to -3.1] | -2.5 ± 0.2 [-2.9 to -2.2] |

**Supplemental Table 4. Group parameters for modeled acetylcholine dose-response curves.** Data are means ± SE [95% Confidence Interval]. See Analytical Procedures for dose-response curve modeling analysis. L-NAME, N^G^-nitro-l-arginine methyl ester.

|  | **Calculated NO-dependent dilation** | **N (M/F)** |
| --- | --- | --- |
| **∆AUC (∆a.u.)** | 147 ± 113 | 67 (33/34) |
| **%AUC (∆%a.u.)** | 47 ± 22 | 67 (33/34) |
| **logEC_50_ (%NOpredicted)** | 28 ± 13 | 69 (34/35) |
| **logEC_90_ (%NOpredicted)** | 40 ± 21 | 76 (38/38) |
| **∆Peak CVC (∆%max)** | 32 ± 20 | 72 (37/35) |

**Supplemental Table 5. Quantification of acetylcholine-induced nitric oxide (NO)-dependent dilation.** Data are means ± SD. Some data were unable to be quantified if the magnitude of the response at the nitric oxide synthase inhibited site was greater than the response at the control site, resulting in a negative value for NO-dependent dilation. In these instances, the data were excluded from the analysis. Final sample size for each calculation is presented in the table. a.u., arbitrary units; AUC, area under the curve; CVC, cutaneous vascular conductance; L-NAME, N^G^-nitro-l-arginine methyl ester.

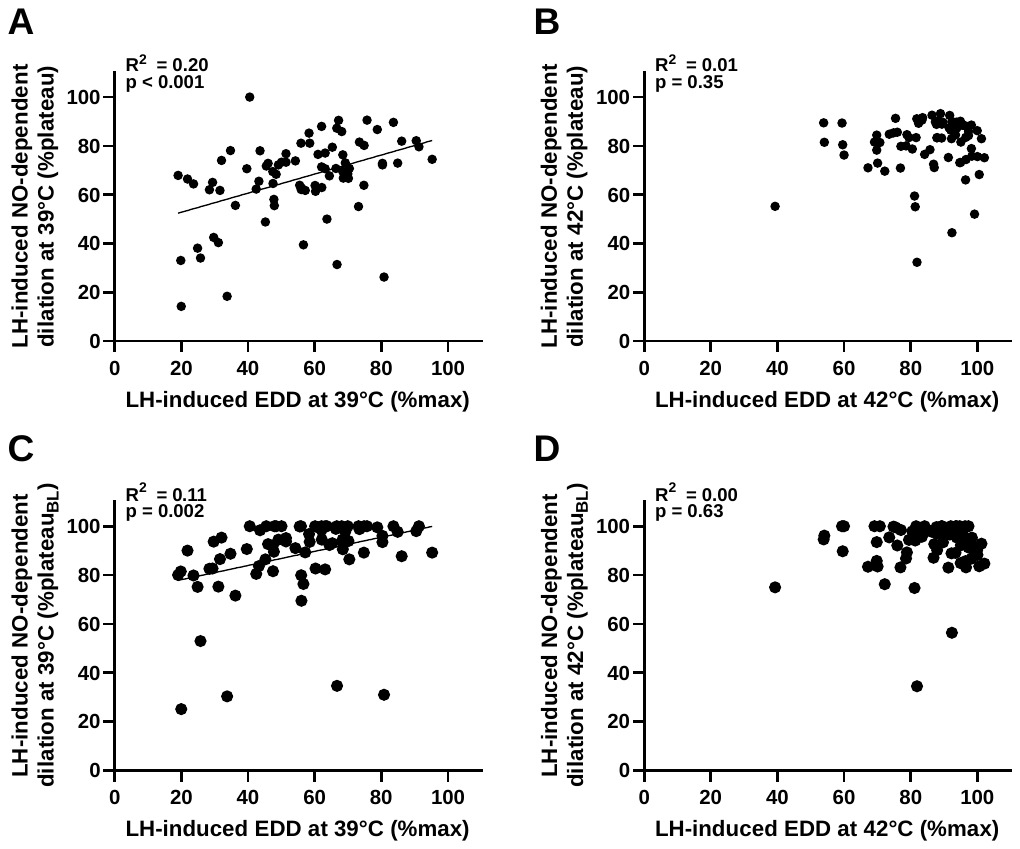

**Supplemental Figure 1.** **Relation between the magnitude of local heating (LH)-induced endothelium-dependent dilation (EDD) to the nitric oxide (NO)-mediated component of that response at 39°C and 42°C.** Associations presented within local heating protocol for 39°C (A and C) and 42°C (B and D). LH-induced NO-dependent dilation at each temperature is expressed as a percentage of the local heating plateau (A and B; %plateau), and as a percentage of the local heating plateau above baseline (C and D; %plateau_BL_). See *Analytical Procedures* for full information of how each index of NO was calculated. Data were analyzed by simple linear regression.
